# Safety and Pharmacokinetics of Intranasally Administered Heparin

**DOI:** 10.1101/2021.07.05.21259936

**Authors:** Hannah M. Harris, Katherine L. Boyet, Hao Liu, Rohini Dwivedi, Nicole M. Ashpole, Ritesh Tandon, Gene L. Bidwell, Zhi Cheng, Lauren A. Fassero, Christian S. Yu, Vitor H. Pomin, Dipanwita Mitra, Kerri A. Harrison, Eric Dahl, Bill J. Gurley, Arun Kumar Kotha, Mahavir Bhupal Chougule, Joshua S. Sharp

## Abstract

**Purpose:** Intranasally administered unfractionated heparin (UFH) and other sulfated polysaccharides are potential prophylactics for COVID-19. The purpose of this research was to measure the safety and pharmacokinetics of clearance of intranasally administered UFH solution from the nasal cavity.

**Methods:** Double-blinded daily intranasal dosing in C57Bl6 mice with four doses (60 ng to 60 μg) of UFH was carried out for fourteen consecutive days, with both blood coagulation measurements and subject adverse event monitoring. The pharmacokinetics of fluorescent-labeled UFH clearance from the nasal cavity were measured in mice by *in vivo* imaging. Intranasal UFH at 2000 U/day solution with nasal spray device was tested for safety in a small number of healthy human subjects.

**Results:** UFH showed no evidence of toxicity in mice at any dose measured. No significant changes were observed in activated partial thromboplastin time (aPTT), platelet count, or frequency of minor irritant events over vehicle-only control. Human subjects showed no significant changes in aPTT time, international normalized ratio (INR), or platelet count over baseline measurements.

No serious adverse events were observed. *In vivo* imaging in a mouse model showed a single phase clearance of UFH from the nasal cavity. After 12 hours, 3.2% of the administered UFH remained in the nasal cavity, decaying to background levels by 48 hours.

**Conclusions:** UFH showed no toxic effects for extended daily intranasal dosing in mice as well as humans. The clearance kinetics of intranasal heparin solution from the nasal cavity indicates potentially protective levels for up to 12 hours after dosing.

## Introduction

The coronavirus disease 2019 (COVID-19) pandemic caused by the severe acute respiratory syndrome-related coronavirus 2 (SARS-CoV-2) has led to severe social and economic consequences spanning the entire world (1, 2). Despite active vaccination programs and implementation of social distancing parameters, the number of infections and deaths in the world remain significantly high and new highly transmissible strains are emerging (3). SARS-CoV-2 is a zoonotic betacoronavirus (4) transmitted via person to person mainly through air droplets (5) and sometimes fecal-oral routes (6), potentially infecting nasal, lung, and intestinal tissues (7). Currently, there are no highly effective therapeutics to treat COVID-19, making effective prevention an essential public health goal.

SARS-CoV-2 entry into host cells is initiated by molecular interaction between the SARS-CoV-2 spike glycoproteins (also known as S-protein) and heparan sulfate proteoglycans (HSPG) displayed on the host cell surfaces (8-11). Several viruses from different virus families are known to utilize HSPG as their attachment receptors (12), for example, parainfluenza (13), adenovirus (14-16), human immunodeficiency virus (HIV) (17, 18), cytomegalovirus (16, 19), and herpes simplex virus (20). In case of SARS-CoV-2, the S-protein-HSPG interaction is followed by binding of S-protein to the human angiotensin converting enzyme 2 (ACE2) (21-26). Binding of SARS-CoV-2 to host HSPG is essential for the progression of virus entry and infection cycle, as evidenced by disruption of virus entry by exogenously administered sulfated polysaccharides, which compete with cell surface HSPGs. The sulfated polysaccharides that have already been investigated as anti-SARS-CoV-2 agents include heparin, heparin derivatives and glycosaminoglycan analogs (9, 10, 27). These sulfated polysaccharides are likely to remain effective against emerging variants of concern (VOC) that partially evade vaccine-induced immune response due to differences in polysaccharide and antibody binding sites on SARS-CoV-2 spike glycoprotein (10) and the conserved nature of the polybasic furin cleavage site, one of the major sites of heparin interaction and activity (28).

Since the major route of virus transmission of SARS-CoV-2 is predominantly initiated by airborne droplet infection of nasal tissues, it is reasonable to consider the design of potentially novel anti-SARS-CoV-2 agents, like UFH, for intranasal administration (9, 11). Such an approach may lead to the discovery of novel anticoronaviral agents with high efficacy, fast mechanisms of action, wide therapeutic windows, and non-invasive self-administration protocols for a possible prophylactic use. While heparin administration by injection is already FDA-approved for other uses, potential antiviral utility against coronaviruses is just being explored.

Unfortunately, little is known about the effects of UFH, particularly regarding the potential consequences of repeated administration to the nasal cavity. Preliminary tests regarding clearance, pharmacokinetics, and safety must be undertaken prior to implementing the actual clinical nasal application of a potentially useful anti-SARS-CoV-2 drug, like heparin, which is already FDA-approved for use administration via other routes. In this work, a series of *in vivo* pharmacological and toxicological aspects of intranasally administered UFH in mice were investigated. Blood histology, coagulation, olfactory behavior, drug clearance, and adverse outcomes were assessed in mice, and safety was evaluated in a small healthy human sample population. Primary objectives for the human study were the measurement of systemic bioavailability based on increased blood coagulation properties using aPTT and INR measurements. Platelet count was also observed to detect any potential heparin-induced thrombocytopenia. Finally, episodes of epistaxis were monitored to measure for localized damage to mucus membranes and/or localized anti-coagulation activity.

Based on the above findings, we hypothesized that the intranasal administration of UFH solution will readily deliver the UFH to the nasal passages and will be safe with no or minimal toxic effects in mice or humans. Results indicate efficient delivery, extended retention of heparin in the nasal passages, and clearance of intranasally administered UFH with no major adverse effects.

## Methods

### Materials

Heparin Sodium Injection, USP was purchased in two different formulations for human intranasal administration: 5,000 USP units per mL (NDC: 71288-403-10) and 10,000 USP units per mL (NDC: 00409-2721-30). Heparin sodium salt from porcine intestinal mucosa (≥ 180 U/mg) was obtained from Sigma-Aldrich (St. Louis, MO) and used without further purification. CF680R dye was obtained from Biotium (Fremont, CA). Reagents for aPTT analysis were obtained from Pacific Hemostasis. aPTT times were measured on a KC4 coagulometer (Sigma Amelung). Other chemicals were purchased from VWR International (Radnor, PA). The U line S-20037C – 1 Oz Glass Dropper Bottles with pump (100 μl per spray) was provided by Brister’s H & W Compounding Pharmacy, Grenada, MS.

### Preparation of Heparin product

All formulation steps were performed under a Biosafety cabinet hood in BSL-2 level laboratory. For animal studies, diluent medium was prepared using 39.6 mL of sterile milliQ water, 0.4 mL benzyl alcohol, and 0.36 g of NaCl. The pH of the diluent was adjusted to 6.2, and the diluent solution was filtered using a 0.2 μm filter. A stock solution of 5 mg/mL of heparin was prepared by dissolving 10.5 mg of dry heparin sodium in 2 mL of diluent solution. The stock solution was then serially diluted 10x using the diluent medium to prepare specified doses of heparin.

For human studies, heparin sodium injection at either 5000U/mL (NDC: 71288-402-10) or 10000U/mL (NDC: 00406-2721-30) was transferred into glass intranasal U line S-20037C – 1 Oz Glass Dropper spray bottles with a 100 μL actuator (Aptar Pharma, Congers NY) under sterile conditions. The intranasal spray bottles were verified to dispense 0.1 mL of heparin solution per pump.

### Animals

All procedures were approved by the University of Mississippi Institutional Animal Care and Use Committee (protocol #19-009) or the University of Mississippi Medical Center Institutional Animal Care and Use Committee (protocol #1571). Toxicology experiments were performed on wild-type male (19-23g) and female (16-19g) C57BL6 mice from Envigo (Indianapolis, IN) in study groups of six mice for each dose (three male, three female). The study was powered to preserve an 80% chance of not committing a Type II error with a 5% chance of Type I error at significant effect sizes (weight as the variable: power > 0.8 to detect a 20% change in body weight within sex (n = 3), 10% change in body weight within entire group (n = 6); aPTT time: power > 0.8 to detect a 100% change in aPTT (n = 4); buried food recovery time: power > 0.8 to detect a 100% change in recovery time (n = 5)). No data were excluded from analysis.

*In vivo* imaging experiments were performed on six (three male, three female) SKH1-Elite mice (Crl:SKH1*-Hr*^*hr*^ Outbred: Charles River Laboratories, Wilmington, MA). One female mouse exhibited poor response to the anesthetic resulting in significant movement during imaging, so data for this mouse were excluded from the nasal clearance kinetics modeling. All animals for toxicology studies were housed in the AALAC-accredited University of Mississippi Animal Facility and given access to rodent chow (Cat.# 7001, Envigo Teklad 4% Fat Rodent Diet) and tap water ad libitum for the duration of the experiments. Mice were group-housed (3-4 mice/cage) in climate-controlled rooms (30-40% relative humidity, 21-23 °C) under a 12:12 h light/dark cycle (lights on at 06:00). All mice were acclimated to the vivarium colony room for five days prior to initiating the drug administration regimen. Upon completion of the study, mice were anesthetized with vaporized isofluorane, perfused with phosphate buffer saline, and secondarily euthanized with decapitation. Tissues of interest were examined, weighed, and fixed in buffered 4% formalin. The left liver lobes were embedded in 30% sucrose (Sigma, S9378) overnight at 4 °C and frozen in optimal cutting temperature for cryosectioning (Leica CM 3050S) at -15 °C. Serial slices (10μm) were transferred to superfrost coated glass slides and stained following standard H&E protocols (dehydrated with alcohol, stained in hematoxylin for 2 minutes, rinsed, differentiated in 70% alcohol and then subsequently stained in 0.01% eosin Y for 30 seconds, rinsed in 95% ethanol, dehydrated with absolute ethanol and cleared in three xylene washes). Entire hepatic cross-sections were imaged (40x) and stitched (10% overlap) using an HCA Nikon Ti2-E microscope and assessed by a blinded observer.

### Intranasal (IN) drug delivery

Mice were transferred to a surgical suite each day and weighed prior to inhaled anesthesia induction with up to 5% isofluorane. Mice were then administered 6 μL per nostril of Heparin solution at concentrations from 5 to 5000 mg/L using a micropipette timed to inspiration. Mice were returned to their home cage and were monitored during recovery (less than 5 minutes). Drug was administered each morning (08:00-12:00) for 14 consecutive days. Signs of inflammation, bleeding, and/or nasal discharge were monitored twice daily by visual inspection of the nose, body, cage, and bedding. Images of the nose were taken prior to administration, one week into administration, and on the 14^th^ day of administration for further visual analyses of inflammation or irritation.

### Olfaction assessment

The Buried Food Test was used to assess the ability to smell volatile odors following the 14-day regimen of IN administration. Mice were habituated to the food stimulus of choice, Teddy Grahams (Nabisco, Hanover, New Jersey), at least 24-hours prior to behavioral testing to confirm its rewarding stimulus. Mice were food deprived (water ad lib) immediately following Day 14 of IN drug administration to motivate subjects, and tested 24-hours later (signs of distress were monitored while food deprived). On test day, the subject was placed in a test chamber containing 3 cm of fresh 1/8 in corncob bedding for a 15-minute trial, with the food stimulus buried approximately 1 cm below the bedding surface in a randomly assigned corner of the cage. The subject successfully completed the assay when it uncovered the food stimulus and began to eat, using one or both forepaws on the food. Latency to uncover and eat the food served as the dependent measure. Each trial had a maximum latency of 900 seconds. Immediately following testing, mice were returned to their home cage, and ad lib access to food was restored.

### Blood histology

At the time of exsanguination, 20 μL of whole blood was removed and smeared on coverglass for histological assessment. The smears were air dried and stained with Wright’s Blood Stain (Fisher, #9513306), washed 3 times, and dried for microscopic imaging. Only the highest heparin dose was analyzed for platelet count. Five fields per sample were imaged at random at 600x phase contrast using a Nikon Ti2-E Microscope. Platelets in each image were counted by a blinded observer.

### aPTT Measurements

Activated partial thromboplastin time assay (aPTT) was performed by incubating 50 μL of test animal plasma with 50 μL of control reference plasma. Following this, 100 μL of aPTT reagent was added and the individual mixtures were further incubated for five minutes at 37 °C. Clotting time was measured in seconds immediately following the addition of 100 μL of 25 mM CaCl_2_. For control values, heparin was added directly to 100 μL of reference plasma at a defined concentration (U/mL), aPTT reagent was added and the aPTT assay performed as described.

### In vivo Imaging of Intranasal Heparin product

CF680R-heparin was generated by conjugating the free reducing-end of heparin with the aminooxy group of the CF680R. Briefly, 50 μM of heparin (EMD Millipore) stock solution was prepared in 100 mM sodium phosphate and 1.5 M sodium chloride (pH 7.4). 5mM of CF680R (Biotium) stock solution was prepared in water. 150 μL of 5 mM CF680R was added to 300 μL of 50 μM heparin. The molar ratio of heparin and CF680R was 1:50. 45 μL of aniline was added to catalyze the reaction. The mixture was reacted at room temperature overnight in the dark. The conjugate was purified by a 3kDa Amicon Ultra centrifugal filter (Millipore Sigma) at 13,000xg for 30 minutes, followed by three washes with 400 μL of water at 13,000xg for 30 minutes. The final sample was further purified by HPLC size exclusion chromatography using a 4.6 mm x 300 mm ACQUITY BEH SEC column (Waters). Sample was analyzed using an isocratic gradient of 40 mM sodium acetate in 20% methanol at a flow rate of 100 μL/min and detection by UV absorbance at 680 nm.

CF680R-heparin (excitation maximum = 680 nm; emission maximum = 701 nm) was resuspended in 1X PBS (600ug/72 μl) and administered at 1 μl per nostril (1.67 μg total) in the mouse anesthetized with 3% isoflurane initially and maintaining anesthesia with 1% isoflurane. Images were acquired on an IVIS Spectrum animal imager system using Living Image® Software (Perkin Elmer). Images were captured before administration of heparin and at 5 min, 15 min, 30 min, 1 h, 2h, 5 h, 12h, 24 h, 48 h, 72 h and 96 h post administration. Quantification was performed by manual definition of the nasal cavity followed by software quantification of IR fluorescence average radiant efficiency in the defined area. Background IR intensity was obtained pre-dosing for each animal and subtracted from each time point. Each replicate was normalized to the intensity of nasal IR fluorescence measured five minutes post-dose for each animal. One-phase and two-phase exponential decay modeling was carried out using GraphPad Prism 9.3.1 and hypothesis testing using an extra sum-of-squares F test was performed. As the *p*-value for the comparison of fits measurement was not below 0.05 (*p* = 0.37) and gains in the goodness of fit parameters were minimal for the two-phase decay model (*R*^2^ = .77 for the one-phase model versus .78 for the two-phase model), the simpler model was chosen.

### Statistical Analysis

Prior to the initiation of treatment, all mice were thoroughly examined for abnormalities and one mouse with a blunted nose was removed and replaced. Drug was assigned to the mice using block randomization for equal group size and drug doses were coded to ensure administration and outcome assessments were performed double blinded. Data were analyzed using SPSS and SyStat SigmaPlot v13 software. Normality was assessed with Shapiro-Wilk test and equal variance was assessed with Brown-Forsythe test. One-way ANOVAs with post-hoc Bonferonni tests were utilized to compare differences with a statistically significant cut-off value of p<0.05. Two-way ANOVAs for sex and treatment revealed a significant main effect of sex across all body weights and most tissue weights, thus each sex was considered independently.

### Human Study Population

An Early Phase 1 interventional clinical trial (NCT04490239) in a small sample population was performed under the approval and guidance of the University of Mississippi Institutional Review Board (protocol #21-004). Six volunteers (three male, three female) between the ages of 18 and 61 (mean age 31 yr) were included in the non-randomized exploratory study after screening ten participants in Oxford, MS (**Table 1**). The six participants were selected based on subject availability during the preferred study dates; the remaining four were retained as replacement subjects in case of participant withdrawal. Exclusion criteria were allergy to heparin; current use of anti-coagulant or anti-platelet drug therapies or any intranasal medication; known history of anemia, thrombocytopenia or other blood disorder; autoimmune disorders; known history of neurologic/psychiatric disorders; reports of any active infection; or subject is pregnant, breast-feeding or is expecting to conceive during the study. All female volunteers were tested for pregnancy via HCG test during each blood draw; no tests returned positive for pregnancy at any point during the study. All subjects submitted written informed consent prior to enrollment in the study. At baseline, all volunteers had clinically normal aPTT (24-33 sec) and INR values (0.8-1.2), along with clinically normal platelet counts (150000-450000 platelets/μL). All participants completed the entire course of the study. This study was not randomized, as all participants received the intranasal heparin intervention.

**Table 1:**
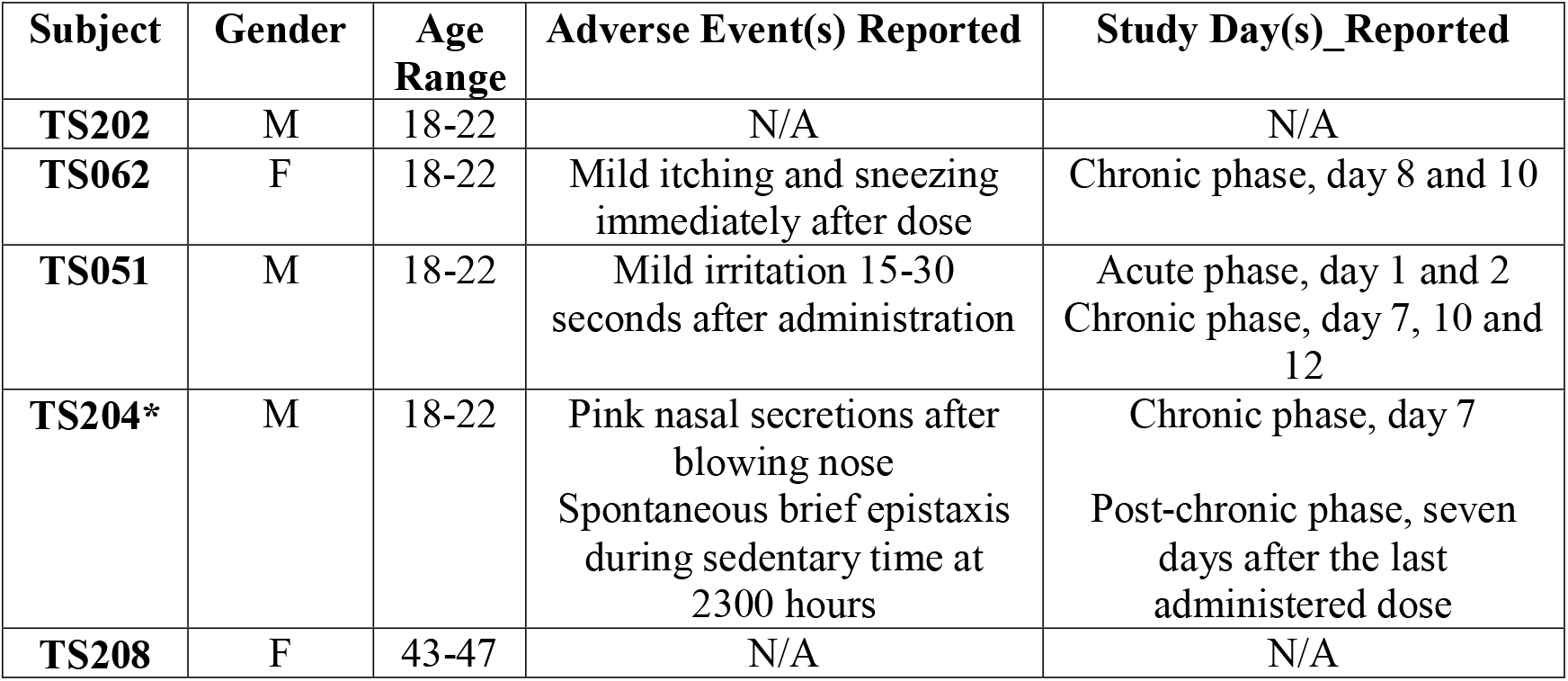
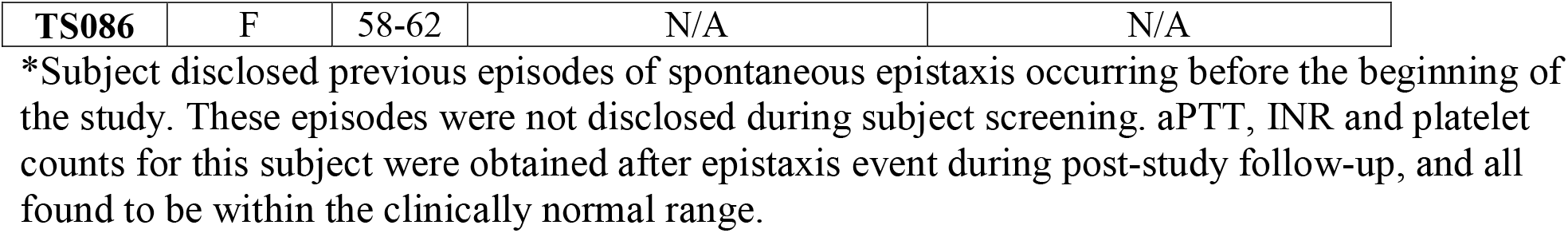
Adverse events observed during the tolerability study of intranasal heparin in human volunteers.

| Subject | Gender | Age Range | Adverse Event(s) Reported | Study Day(s)_Reported |
| --- | --- | --- | --- | --- |
| TS202 | M | 18-22 | N/A | N/A |
| TS062 | F | 18-22 | Mild itching and sneezing immediately after dose | Chronic phase, day 8 and 10 |
| TS051 | M | 18-22 | Mild irritation 15-30 seconds after administration | Acute phase, day 1 and 2<br>Chronic phase, day 7, 10 and 12 |
| TS204* | M | 18-22 | Pink nasal secretions after blowing nose<br>Spontaneous brief epistaxis during sedentary time at 2300 hours | Chronic phase, day 7<br>Post-chronic phase, seven days after the last administered dose |
| TS208 | F | 43-47 | N/A | N/A |
| **TS086** | F | 58-62 | N/A | N/A |

\*Subject disclosed previous episodes of spontaneous epistaxis occurring before the beginning of the study. These episodes were not disclosed during subject screening. aPTT, INR and platelet counts for this subject were obtained after epistaxis event during post-study follow-up, and all found to be within the clinically normal range.

### Administration of Intranasal Heparin to Human Volunteers

Administration was carried out in two phases: an acute phase to determine tolerability of two doses, and a chronic phase to determine tolerability of repeated daily doses using U-line nasal spray bottle with pump over a two-week period. During the acute phase, volunteers applied a single spray of 500U to each nostril on Day 1 in the clinic and volunteers were observed for one hour for adverse events. Whole blood was drawn by venipuncture 24 hours after dosing to allow for aPTT measurement. If the aPTT value remained in the clinically normal range, volunteers applied a single spray of 1000U to each nostril on Day 3 in the clinic. Again, aPTT was measured 24 hours post-dosing. No differences in aPTT levels were observed at either dose in the acute phase, thus all participants entered the chronic phase of the intervention. A five-day washout period was used between the acute phase and the chronic phase.

During the chronic phase, each volunteer had a single 1000U heparin spray per nostril administered on Day 1 in the clinic, and the subject was observed for adverse events. Subjects were instructed on how to self-administer the heparin dose, and subjects were given the test article to self-administer daily at home. Volunteers maintained a dosing journal to document self-administration. Follow-up calls were made on Day 4, Day 7, Day 9 and Day 11 to check on subject compliance, response and adverse events. Blood was drawn after the final dose on Day 14 in the clinic for aPTT measurement. Blood was drawn 24 hours after the final dose, on Day 15, and two weeks following completion of the study (Day 30) for aPTT, PT/INR and platelet count measurements. All hematological assays were performed by a contract medical testing lab. Results were registered with clinicaltrials.gov (NCT04490239). No changes were made to methods or assessment criteria after trial commencement.

## Results and Discussion

### Chronic Toxicology of Intranasal Heparin in a Mouse Model

Varying doses of heparin or vehicle control were administered to the nasal cavities of male and female mice once per day for 14 consecutive days to monitor safety and tolerability with this route of administration. Block randomization was used to assign dosing groups of 5 mg/L (0.06 μg/day), 50 mg/L (0.6 μg/day), 500 mg/L (6 μg/day), and 5000 mg/L (60 μg/day) heparin formulated in 5 mg/mL sodium chloride, 1% benzyl alcohol (pH 5.0 – 7.5). 12 μL (6 μL/nostril) were administered daily via drop pipetting into the nasal cavity during inspiration. Notes were taken on animal appearance, grooming, signs of distress, and other notable behaviors twice daily throughout the treatment period. Detailed results are shown in **Supplementary Table 1**. As expected with this developmental stage, both cohorts of male and female mice exhibited small weight gains over the 14-day drug treatment. Two-way multiple comparison ANOVA indicated a main effect of sex on weight, but no treatment effect when all groups were compared. When sex was considered independently, a one-way ANOVA revealed no difference in the extent of weight gain between any of the treatment doses in females (p>0.05). In males, between group differences were observed and a post-hoc Bonferonni test revealed a statistically significant increase in weight gain in mice treated with 6 μg/day heparin versus vehicle-only control (mean: 1.33g gain vs 0.33g gain, p=0.018). No differences in the weights of mice receiving lower or higher doses were observed (**Figure 1**).

**Figure 1.**
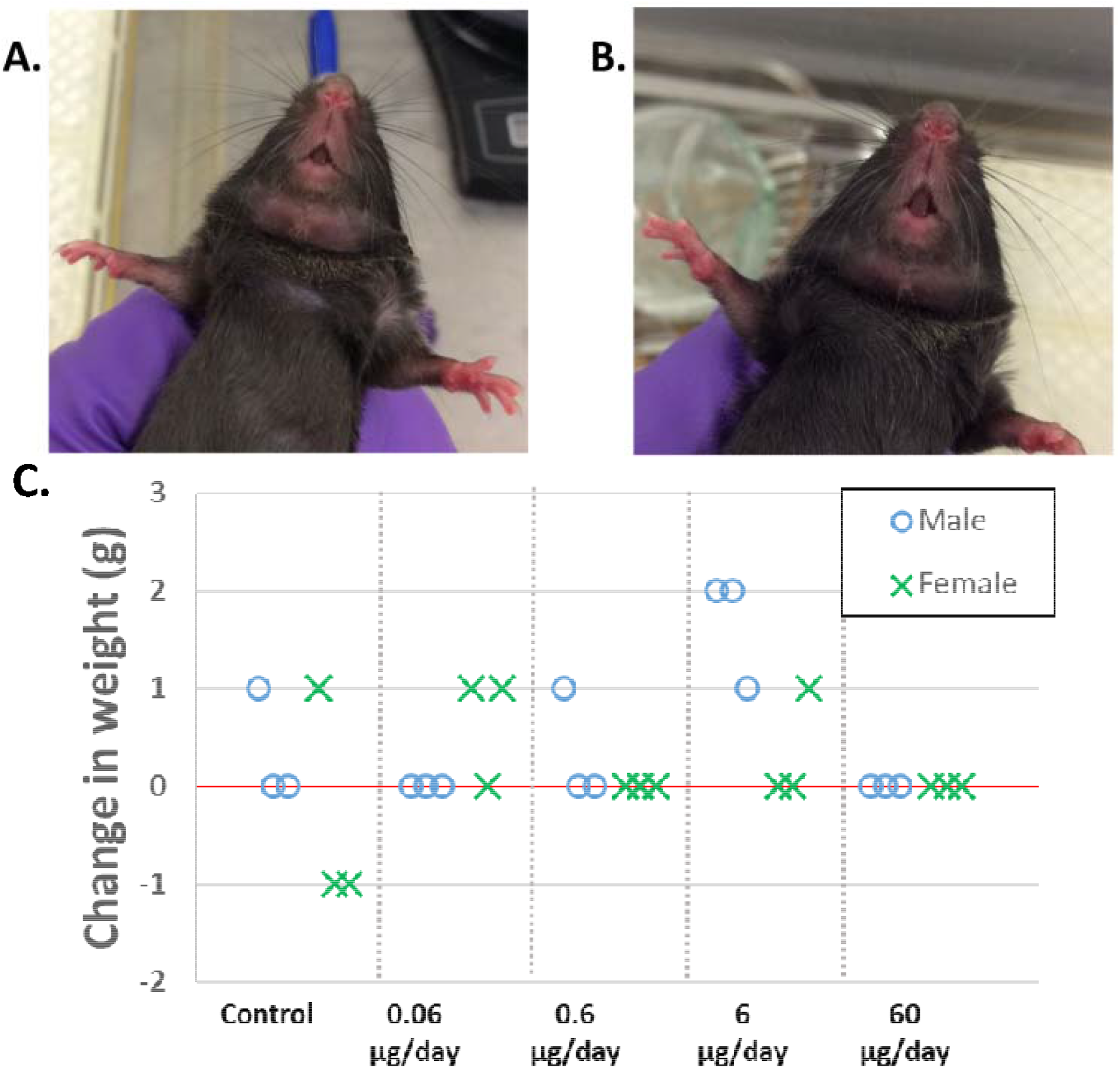
Sample results from 14 day toxicology trial of intranasal heparin in mice. Images of representative **A**. vehicle control and **B**. 60 μg/day mouse nose after two weeks. **C**. Change in body weight from pre-dose to after the 14^th^ day dose.

Minor nasal discharge, minor nose wetness, dried nasal residue, and sneezing or nasal bubbling upon administration were observed in both test and vehicle-only groups. Minor irritation (pink discoloration) and minor swelling of the nose was observed in a few animals throughout the 14 day protocol, with no differences in prevalence across groups. Only one mouse (male, 6 μg/day heparin) showed irritation on more than one day of the study and the three days of observed irritation were non-consecutive and ended on day 10. No signs of epistaxis or topical bleeding were evident in any mice during the study (**Figure 1**). Overall, the frequency of observed minor effects for heparin-dosed animals were small and consistent between males and females (0.29±0.07 events per day for males, 0.29±0.10 events per day for females). No dose-dependence was observed in the frequency of minor effects for heparin-dosed animals (**Figure 2**). The frequency of observed minor effects in vehicle-only control animals in males (0.22±0.10 events per day) or females (0.49±0.14 events per day) was not significantly different from heparin-dosed animals (p>0.05), suggesting the minor irritation may be attributed to the technical aspects of inspiration-timed intranasal administration in young mice.

**Figure 2.**
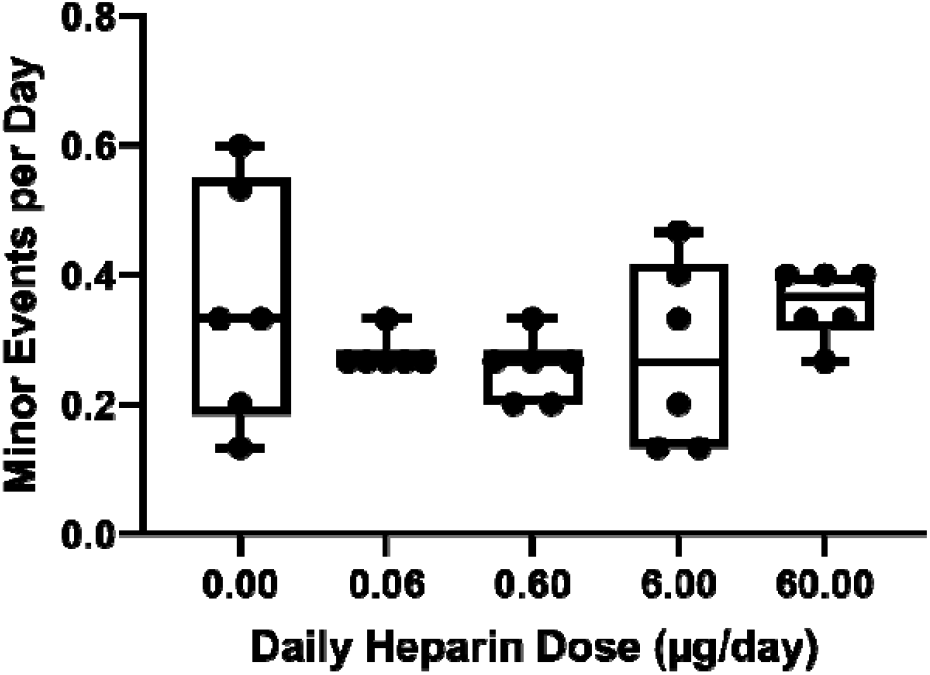
Dose dependence of frequency of minor observations reported for 14-day heparin trial in mice. No dose dependence in the frequency of minor observations (nasal discharge, dried nasal residue, mild nasal discoloration, etc.) was observed, and no difference between heparin-dosed and vehicle-only control animals were observed.

Necropsies were performed at the completion of the drug treatment paradigm with no overt pathology noted across all mice. The wet weights of heart, lungs, liver, spleen, and both kidneys were compared. The only significant difference in wet organ mass observed compared to vehicle-only control was a decrease in liver mass in females receiving the 6 μg/day dose (0.22g decrease; p = 0.0207). When normalized to total body weight, one-way ANOVA analysis with post-hoc Bonferroni test revealed a small but significant decrease in relative liver:body mass ratio weight in females at 0.6 μg/day (0.0127g decrease, p = 0.0009), 6 μg/day (0.0131g decrease, p = 0.0007) and 60 μg/day (0.0112g decrease, p = 0.0022). Histological assessments of the 6 μg/day and control females revealed no evidence of hepatotoxicity or neoplasia. No significant change was found in liver:body mass ratio for any individual dose in males. All necropsy data are presented in **Supplementary Table 1**. Whole blood was sampled upon completion of the 14 day dosing regimen and platelet count at this time point revealed no significant difference across all treatment groups (p = 0.766, **Figure 3**), indicating no signs of heparin-induced thrombocytopenia. In vitro analysis of coagulation time of plasma isolated at this time point also revealed no significant difference in aPTT across all treatment groups (**Figure 4**), indicating no clinically significant systemic bioavailability of daily administered intranasal heparin. As expected, inclusion of 1 U/mL and 0.5 U/mL exogenous heparin in the in vitro aPTT test as a positive control resulted in significant differences (p <0.0001) in aPTT clotting time (**Figure 4**), thereby supporting the efficacy of the aPTT assay.

**Figure 3.**
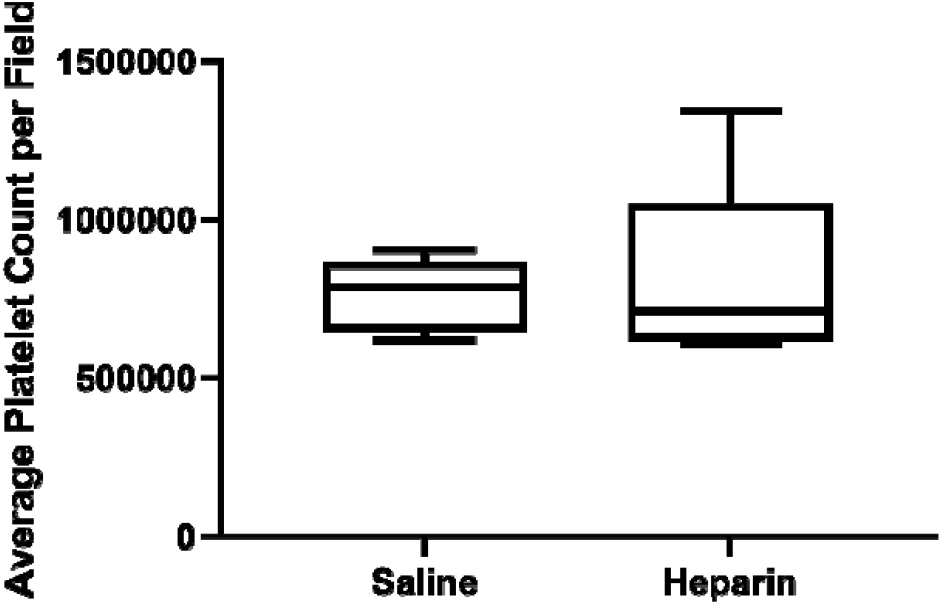
Blood platelet count after 14 days of daily administration of 60 μg of intranasal heparin in mice. No significant difference in platelet count was observed (p = 0.766).

**Figure 4.**
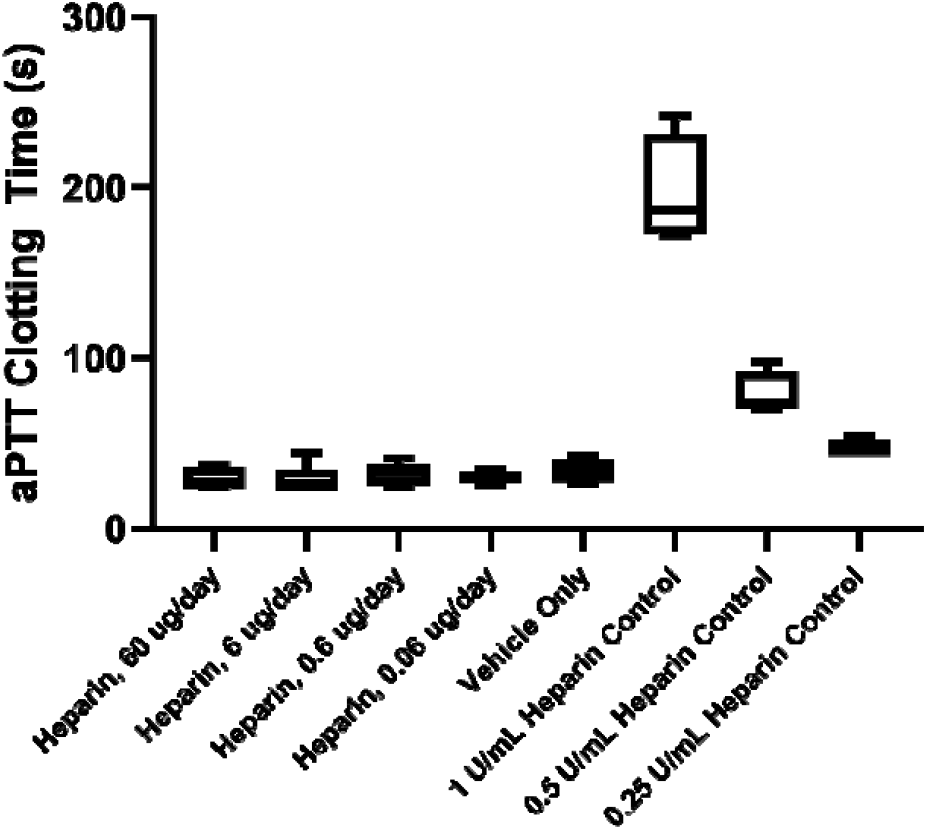
aPTT assay of mouse plasma after 14-day heparin trial. No significant differences in aPTT clotting time was observed between heparin at any dose versus vehicle only. Significant differences were observed between vehicle only and the vehicle plasma with exogenous heparin added directly at concentrations of 1 U/mL or 0.5 U/mL (p < 0.0001).

To assess whether repeated intranasal administration disrupted olfactory function, the buried food test was performed after the final drug administration with the saline and 60 μg/day heparin treatment groups (**Figure 5**). Control naïve mice who did not receive any intranasal treatments were also assessed to determine whether physical administration to the nose each day altered the behavioral phenotype. All mice successfully uncovered the food reward, with no significant difference in the latency to complete the task between any group (one-way ANOVA, p=0.2387). One outlier was observed in the 60 μg/day group with an unusually long time to retrieve the buried food; the subject dug at the location of the food after 43 seconds, but waited to retrieve it. Overall, no evidence of overt toxicity, intolerability, or systemic bioavailability of intranasal heparin was observed in the mice during the two-week dosing period at any dose, up to 60 μg/day.

**Figure 5.**
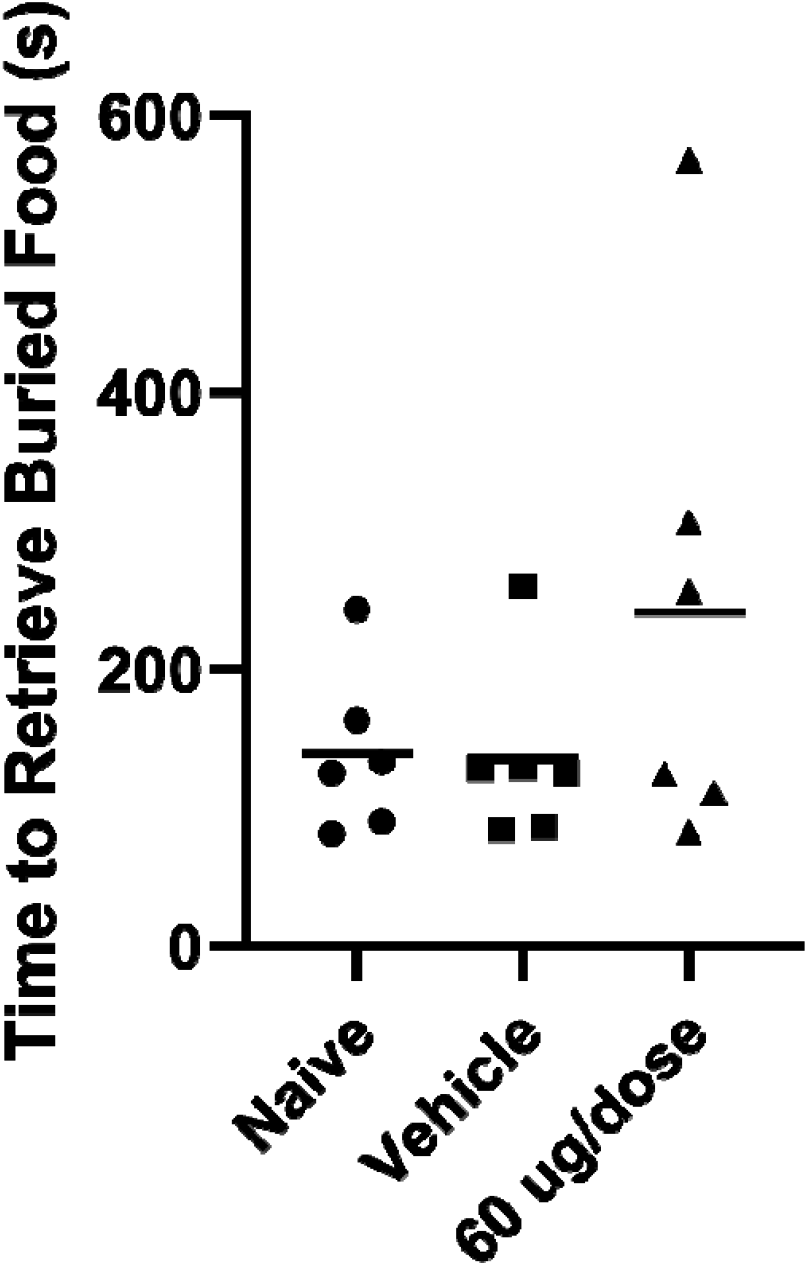
Buried food retrieval anosmia assay of naïve, vehicle-only, and 60 μg/day heparin in mice. No significant differences were observed between groups (p=0.2387).

### In vivo imaging of instranasal heparin clearance kinetics

Of key concern in any therapeutic strategy involving nasal administration of heparin to block viral attachment and entry to nasal epithelial cells is the pharmacokinetics of heparin clearance from the nasal cavity. Unlike residence in the lungs (29), the mucosa of the nasal cavity is constantly cleared into the stomach which can drive intranasal drug clearance. However, heparin has many physical and chemical features that are not common in nasally administered drugs, being much larger and having high negative charge density. To test the lifetime of heparin in the nasal cavity, we labeled heparin at the reducing end with a near-IR chemical dye through an oxime linkage, resulting in a single dye molecule (95 Da) per heparin molecule (avg. ∼15,000 Da). After purification, the resulting CF680R-heparin showed no substantial presence of unconjugated dye by analytical size exclusion chromatography (**Figure 6A**). The resulting CF680R-heparin has biophysical characteristics almost identical to unmodified heparin, while having quantitative near-IR fluorescence that can be imaged in the living mouse. Due to the intensity of the signal, we administered a dose of 1.67 μg of CF680R-heparin to the mouse and quantified the fluorescence signal in the nasal cavity. A comparison of fits test failed to show a statistically significant improvement in model quality for a two-phase model (*p* = 0.37), with a one-phase exponential decay model fitting the observed data well (**Figure 6B**). The observed half-life of the heparin in the nasal cavity was 39.34 min (95% CI 22.7 – 66.6 min). At the goal of 12 hours protection time post-dose, we retain ∼3.2% of the initial heparin dose in the nasal cavity. Based on an average mouse nasal cavity volume of 32 mm^3^ (30) and an initial dose of 60 μg found to be safe for repeated daily dosing in mice, our model indicates we should retain 60 mg/L heparin in the nasal cavity after 12 hours, 10,000 times higher than the heparin IC_50_ measured for inhibiting pseudotyped SARS-CoV-2 attachment and entry (9). Movement of the fluorescence signal is consistent with passage through the nasopharynx and into the digestive tract during mucus clearance (**Figure 6C**).

**Figure 6.**
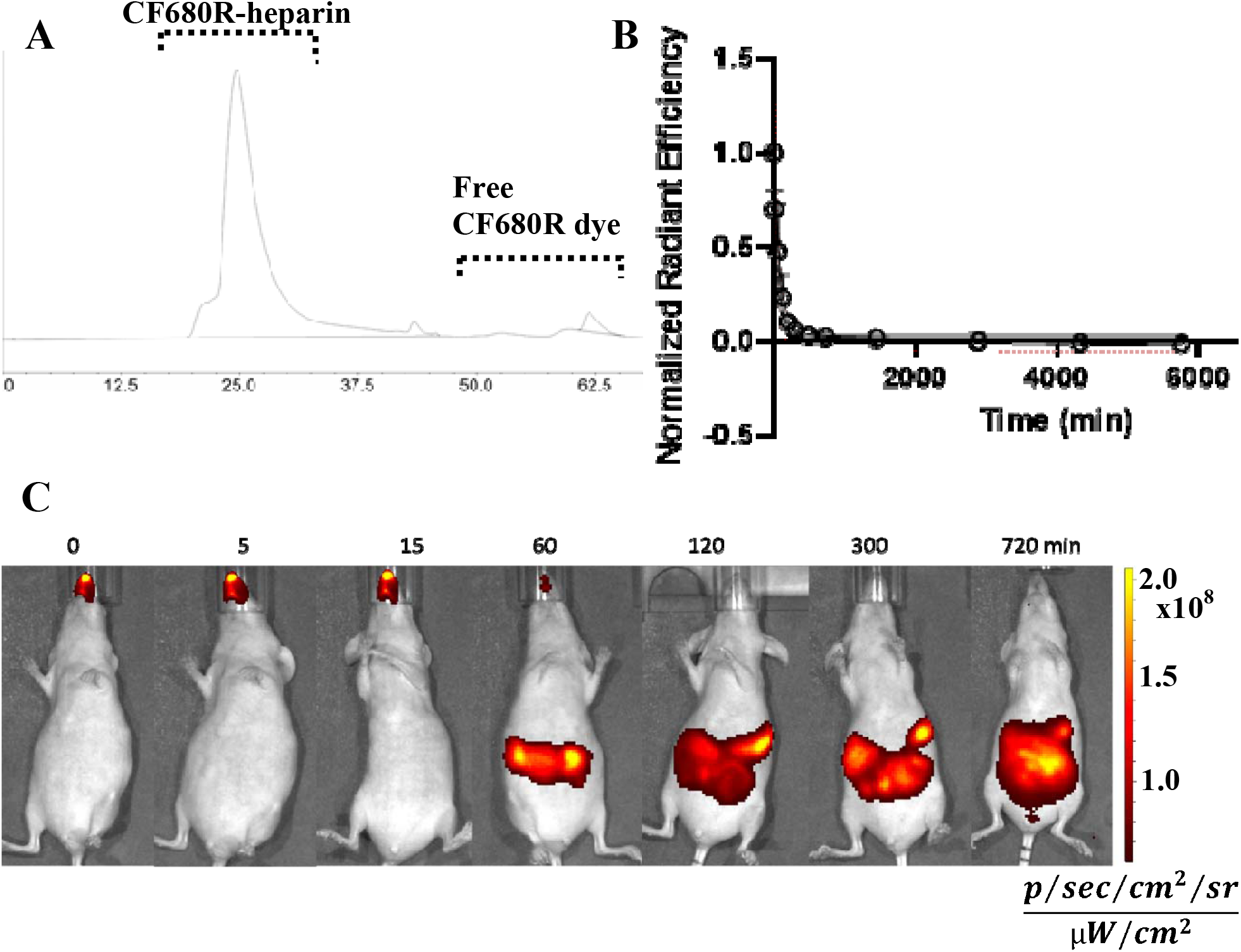
*In vivo* imaging of 1.67 μg of nasally administered CF680R-heparin in mice. **(A)** Analytical size exclusion chromatography verifies that purification has removed nearly all free dye, resulting in all IR fluorescence signal coming from CF680R-heparin. **(B)** Clearance from the nasal cavity follows a one-phase exponential decay (black line; 95% CI shown as red dotted line) to reach background IR levels (normalized to zero); K=0.01762, Y_0_=1.070, half life =39.34 min. Signal is normalized to the measured radiant efficiency at 5 minutes. **(C)** *In vivo* imaging shows accumulation and residence in the nasal cavity, with clear evidence of clearance through the gut.

### Tolerability of intranasal heparin in healthy human volunteers

Heparin was administered to six healthy human volunteers (three male, three female) in two phases. The initial acute phase was performed at two intranasal doses (1000 U/day and 2000 U/day administered as one spray per nostril daily) to monitor for acute adverse effects. No adverse effects were reported during the acute phase, so the chronic phase was performed at 2000 U/day administered as one spray per nostril daily to monitor for longer-term effects after fourteen consecutive days of daily dosing. Effects of intranasal heparin on blood coagulation parameters were followed via an aPTT test 24 hours after each dose in the acute phase, as well as immediately after the final dose in the chronic phase, 24 hours after the final dose in the chronic phase, and 16 days after the final dose in the chronic phase (16 day washout period). An INR test was also performed 24 hours after the final dose in the chronic phase, as well as 16 days after the final dose in the chronic phase. In all cases, no evidence of systemic bioavailability of heparin was observed based on changes in blood coagulation times compared to the pre-study screening results or the clinically normal range (**Figure 7A and B**). Platelet counts were also performed during pre-study screening, immediately after the final dose in the chronic phase, 24 hours after the final dose in the chronic phase, and 16 days after the final dose in the chronic phase. No evidence of heparin-induced thrombocytopenia was observed based on changes in platelet count compared to pre-study values or clinically normal ranges (**Figure 7C**). Subjects also reported mild adverse events, summarized in **Table 1**. Two subjects reported mild nasal irritation and/or sneezing after administration, consistent with exposure to the benzyl alcohol found in the heparin formulation. One subject reported spontaneous brief epistaxis during sedentary time on Day 7 of the chronic phase, as well as seven days after the last dose of the chronic phase. Follow-up questioning indicated that this subject had a history of epistaxis that was undisclosed during screening. These early Phase 1 results support that heparin administered intranasally is tolerableat doses up to 2000 U/day for at least 14 days, with no clinically significant systemic distribution of the heparin and no effect on blood coagulation.

**Figure 7.**
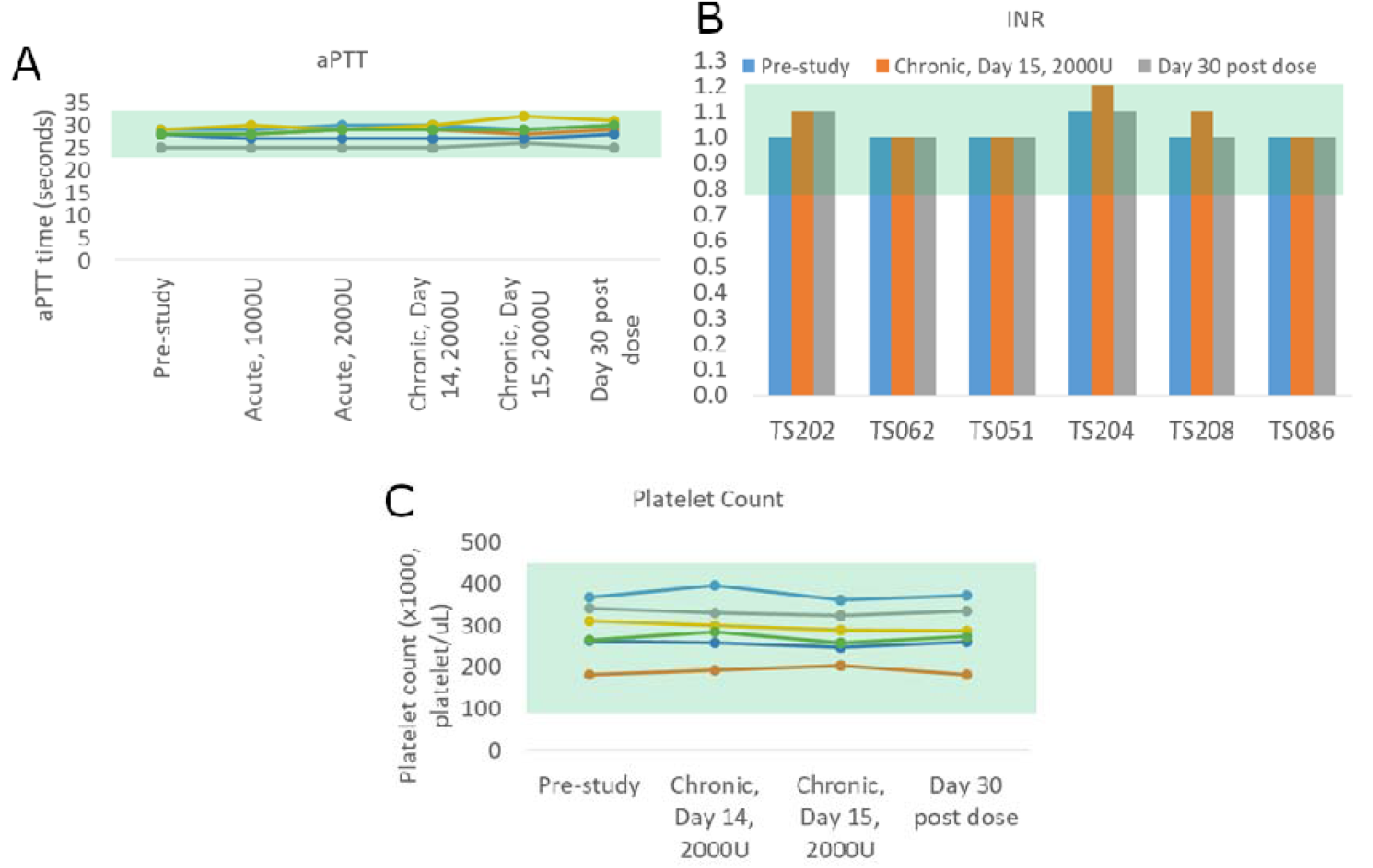
Blood results from intranasal heparin human subjects. Clinically normal range shaded in green. **(A)** aPTT results taken during screening, acute and chronic phases, and wash-out. Each line represents a subject. **(B)** INR results taken during screening, after chronic phase, and after wash-out. **(C)** Platelet count taken during screening, after chronic phase, and after wash-out. Each line represents a subject.

## Conclusion

Our results demonstrated that UFH produces no local nor systemic adverse effects when administered intranasally to either mice or human volunteers. We observed no signs of nasal inflammation, epistaxis, alterations on platelet count that could lead to potential heparin-induced thrombocytopenia, and significant changes on the aPTT analysis. Our findings revealed no overt signs of toxicity nor disruption of olfactory functions in mice administered UFH intranasally over two weeks at doses up to 60 μg/day. From the findings of *in vivo* imaging, we observed a two-phase clearance of intranasal UFH from the nasal cavity, with protective concentrations still present 12 hours post-dosing, in which approximately 2% of the starting dose remained in the cavity until background levels were again reached after 12 additional hours. No significant changes were observed for aPTT, PT/INR, and platelet counts in human volunteers, indicating that intranasal administration of heparin is tolerable at doses up to at least 2000 U/day for at least 14 days. The lack of both local irritation and systemic bioavailability suggest that intranasal heparin may have a high tolerance with a potentially large therapeutic window for preventing viral infections from respiratory viruses that use heparan sulfate as a co-receptor, including SARS-CoV-2.

## Data Availability

The data that support the findings of this study are available from the corresponding author upon reasonable request.

## Acknowledgement

This work was supported in part by the University of Mississippi and the NIH funded Neuropharmacology Core Facility (P30 GM122733). Rodent *in vivo* imaging was performed in the Animal Imaging Core Facility at the University of Mississippi Medical Center. RT is supported by University of Mississippi Medical Center COVID-19 funds and NASA award (80NSSC19K1603). The authors are thankful to Dr. Ward Brister, Brister’s H & W Compounding Pharmacy, Grenada, MS for providing the U line nasal spray bottle with pump.

